# Is Higher Viral Load in SARS-CoV-2 Associated With Death?

**DOI:** 10.1101/2020.08.04.20164061

**Authors:** Klinger Soares Faíco-Filho, Victor Cabelho Passarelli, Nancy Bellei

## Abstract

**Background:** There is no proven prognostic marker or adequate number of studies in patients hospitalized for coronavirus disease 2019 (COVID-19).

**Methods:** We conducted a retrospective cohort study of patients hospitalized with COVID-19 from March 14 to June 17, 2020, at São Paulo Hospital. SARS-CoV-2 viral load was assessed using the cycle threshold (Ct) values obtained from an RT-PCR assay applied to the nasopharyngeal swab samples. Disease severity and patient outcomes were compared.

**Results:** Among the 875 patients, 50.1% (439/875) had mild, 30.4% (266/875) moderate, and 19.5% (170/875) severe disease. A Ct value of <25 (472/875) indicated a high viral load, which was independently associated with mortality (OR: 0,34; 95% CI: 0,217–0,533; *p* < 0.0001).

**Conclusions:** Admission SARS-CoV-2 viral load is an important surrogate biomarker of infectivity and is independently associated with mortality among patients hospitalized with COVID-19.

## INTRODUCTION

The pandemic of severe acute respiratory syndrome coronavirus 2 (SARS-CoV-2) has been responsible for causing >11 million infections around the world since December 2019 [1]. Although the severity is mild in most cases, up to 33% of the infected patients require hospitalization and almost 20% die [2,3]. To date, several studies have focused on the risk factors for moderate to severe coronavirus disease-2019 (COVID-19), such as comorbidities, older age, and laboratory abnormalities [4,5]. However, the effect of SARS-CoV-2 viral load on clinical outcomes remains understated. Two Chinese studies demonstrated that the viral load in hospitalized patients was higher [6,7] and one recent American study demonstrated that higher viral loads were correlated with a higher risk of intubation and death for inpatients [8]. Nevertheless, the correlation between viral load and hospitalization risk in inpatients and outpatients remains unelucidated.

Detection of SARS-CoV-2 by reverse transcription-polymerase chain reaction (RT-PCR) assay in nasopharyngeal swab specimens is the primary diagnostic method for COVID-19 [9]. Data concerning cycle threshold (Ct) values, which are inversely proportional to the amount of RNA viral copies, has been used as an inference of the viral load.

Viral load can also act as a biomarker of disease severity, clinical outcome, and mortality. We conducted a retrospective analysis of SARS-CoV-2 Ct values, disease severity, and clinical outcome of 875 patients at a large university hospital in São Paulo, Brazil, between March and June 2020.

## METHODS

### Study population and setting

This retrospective observational cohort study involved patients who presented with respiratory symptoms from March 14 to June 17, 2020, at São Paulo Hospital in the Federal University of São Paulo, Brazil. A nasopharyngeal swab sample was collected from all of the patients and analyzed for SARS-CoV-2. All the specimens were stored in 2 mL sterile Ringer’s lactate solution in sterile tubes and transported to the Virology Laboratory at the Federal University of São Paulo for testing.

### Viral load analysis method

We used the cycle threshold (Ct) values as a semiquantitative measure of viral load. The amount of viral RNA copies present in the positive samples is inversely proportional to the corresponding Ct value. That is, the greater is the amount of viral RNA, the lower is the Ct value.

### Samples and RNA preparation

Nasal swab samples were collected from the patients at admission. The RNA of the samples was purified using the Quick-RNA Viral Kit (Zymo Research, USA), according to manufacturer’s instructions. The purified RNA was stored at -80°C.

### SARS-CoV-2 detection

Viral detection was performed using the AgPath-ID One-Step RT-PCR reagents (ThermoFisher Scientific, Austin, USA), according to the manufacturer’s instructions. The reactions were performed in a 20 μL total reaction volume containing 5.0 μL purified RNA, 400 nM primers, and 200 nM probes following the CDC USA protocol targeting the N1 and N2 sequences of the SARS-CoV-2 nucleoprotein gene and human ribonuclease P gene (RNAse P) serving as an endogenous control [10]. For the semiquantitative analysis, we used the Ct values of the more sensitive N2 target. Samples with Ct values <40 were considered positive.

### Data collection

Information on age, hospitalization status, and clinical outcome was retrieved from the electronic medical records of the patients. According to World Health Organization guidelines, patients were stratified into groups on the basis of age, as follows: 0–5; 5–14; 15–24; 25–34; 35–44; 45–54; 55–64; 65–74; 75–84; and >85 years. Disease severity and clinical outcome were classified as follows: mild (no hospitalization), moderate (hospitalization in the ward), severe (hospitalization in the Intensive Care Unit), and discharge or death. Patients who were still hospitalized, awaiting a defined outcome, during data collection were not considered for the statistical analysis.

### Statistical analysis

Statistical analyses were performed using the Student *t*-test for parametric data and the Mann–Whitney test for non-parametric data. Significance level was set at a *p* value of <0.05.

## RESULTS

Among the 875 individuals with laboratory-confirmed COVID-19, more than a half (50.9%, 446/875) were female patients. The median age was 48 years (range, 2–97 years) and the median Ct value was 24. A Ct value of <25 (472/875) indicated high viral load and >24 (403/875) low viral load.

The initial Ct values for each age group is presented in Figure 1. We found no significant differences.

**Figure 1:**
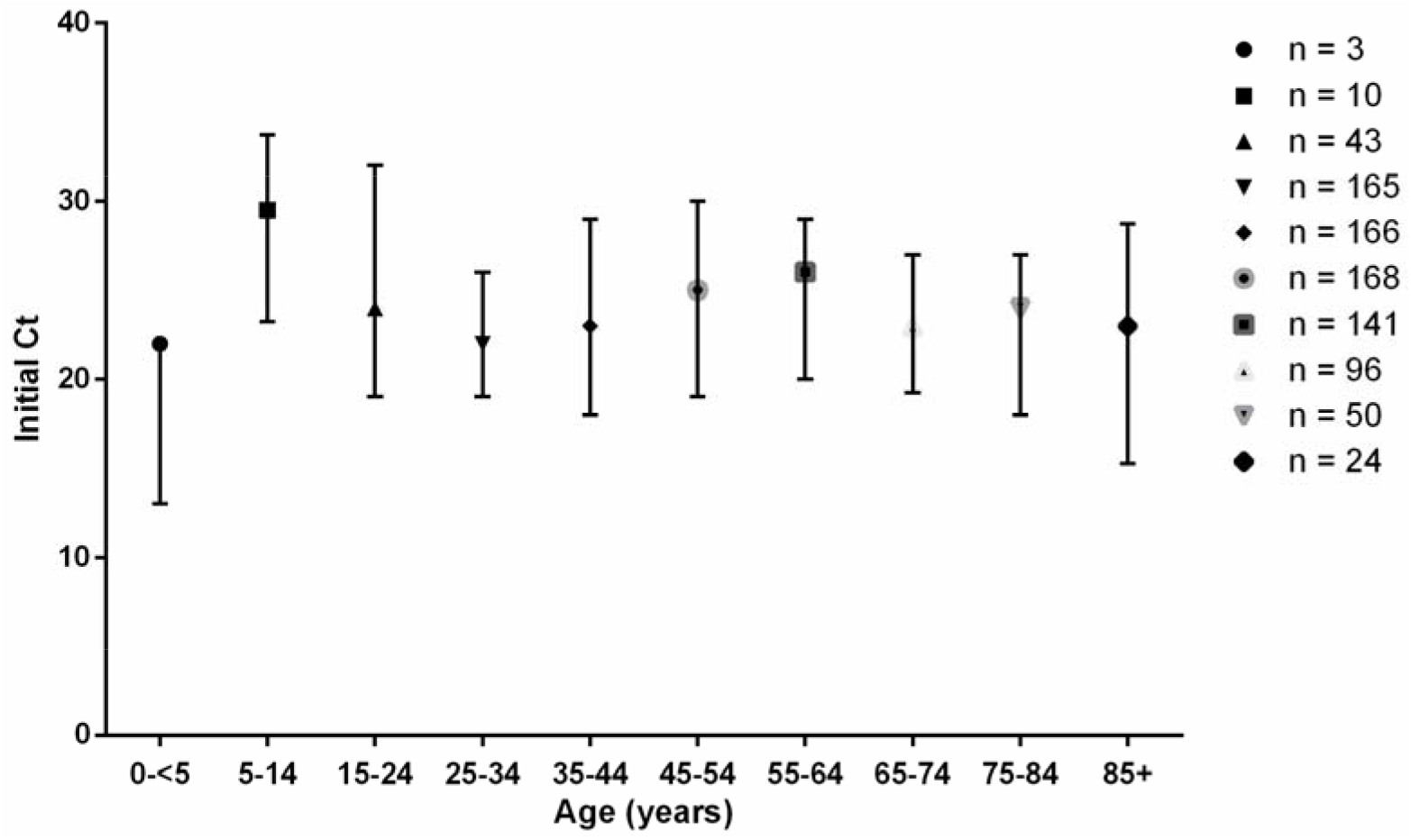
Initial Ct values from the swab samples in different age groups.

The Ct values were analyzed according to the disease severity—mild (50.1%; 439/875; median, 22); moderate (30.4%; 266/875; median, 27), and severe (19.5%; 170/875; median, 21.5)—in the 875 patients. The initial Ct value for patients with moderate disease was higher and significantly differed from that for patients with mild (*p* < 0.0001) and severe (*p* < 0.0001) disease, as is shown in Figure 2.

**Figure 2:**
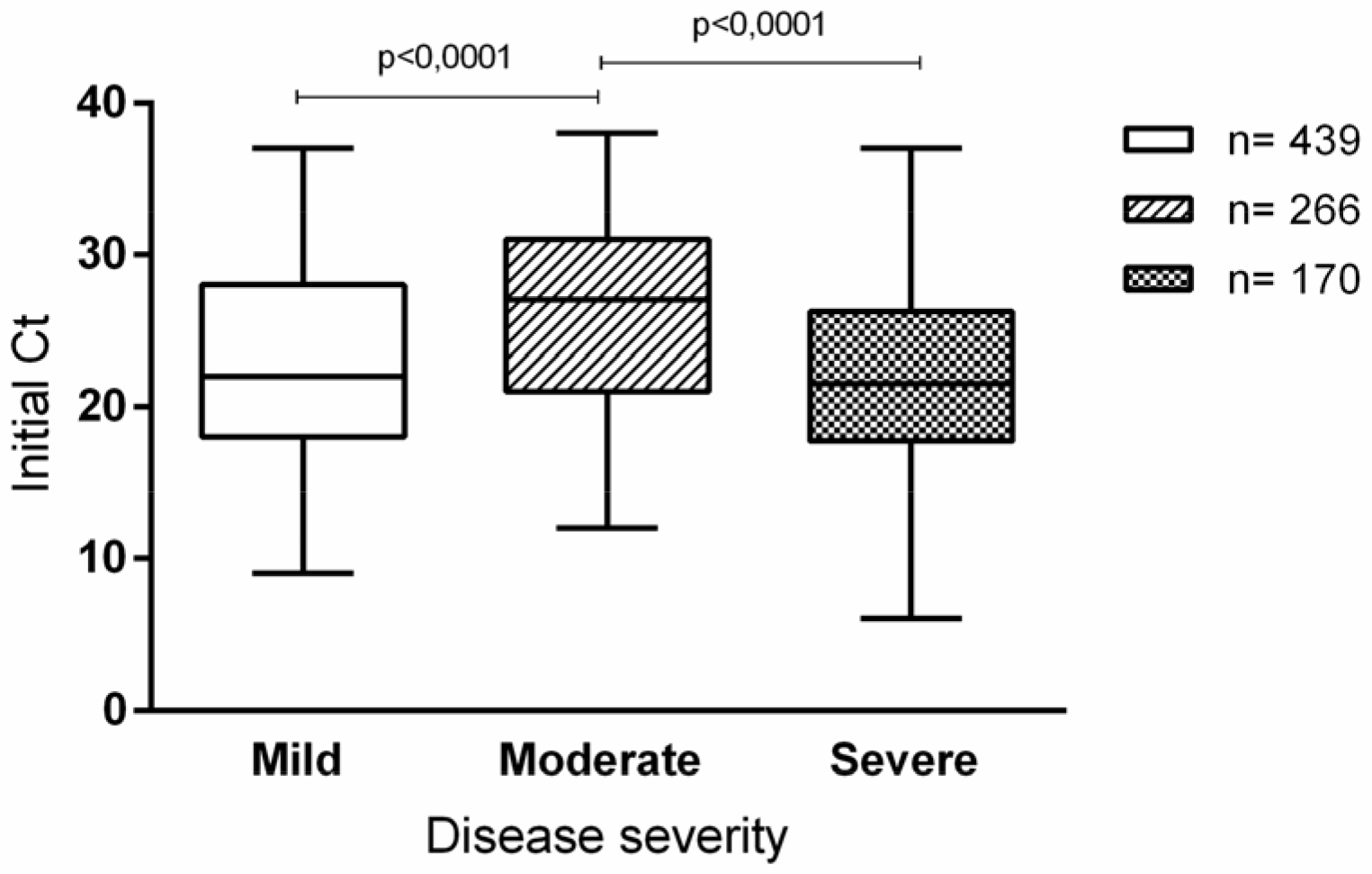
Initial Ct values from the swab samples obtained from patients with different clinical outcomes.

As is shown in Figure 3, when the initial Ct was compared with the hospital discharge or death, survivors demonstrated a higher value (low RNA viral copies) than non-survivors (Ct median, 27 and 21, respectively). The initial Ct differed significantly between the two groups (*p* < 0.0001). Mortality rates were 46% (87/191) among patients with a high viral load (Ct < 25) and 22% (41/185) among patients with a low viral load. The risk of in-hospital mortality was also higher in patients with a high viral load (Ct < 25) than those with a low viral load (Ct > 24), and this association was statistically significant (OR: 0.34; 95% CI: 0.217–0.533; *p* < 0.0001).

**Figure 3:**
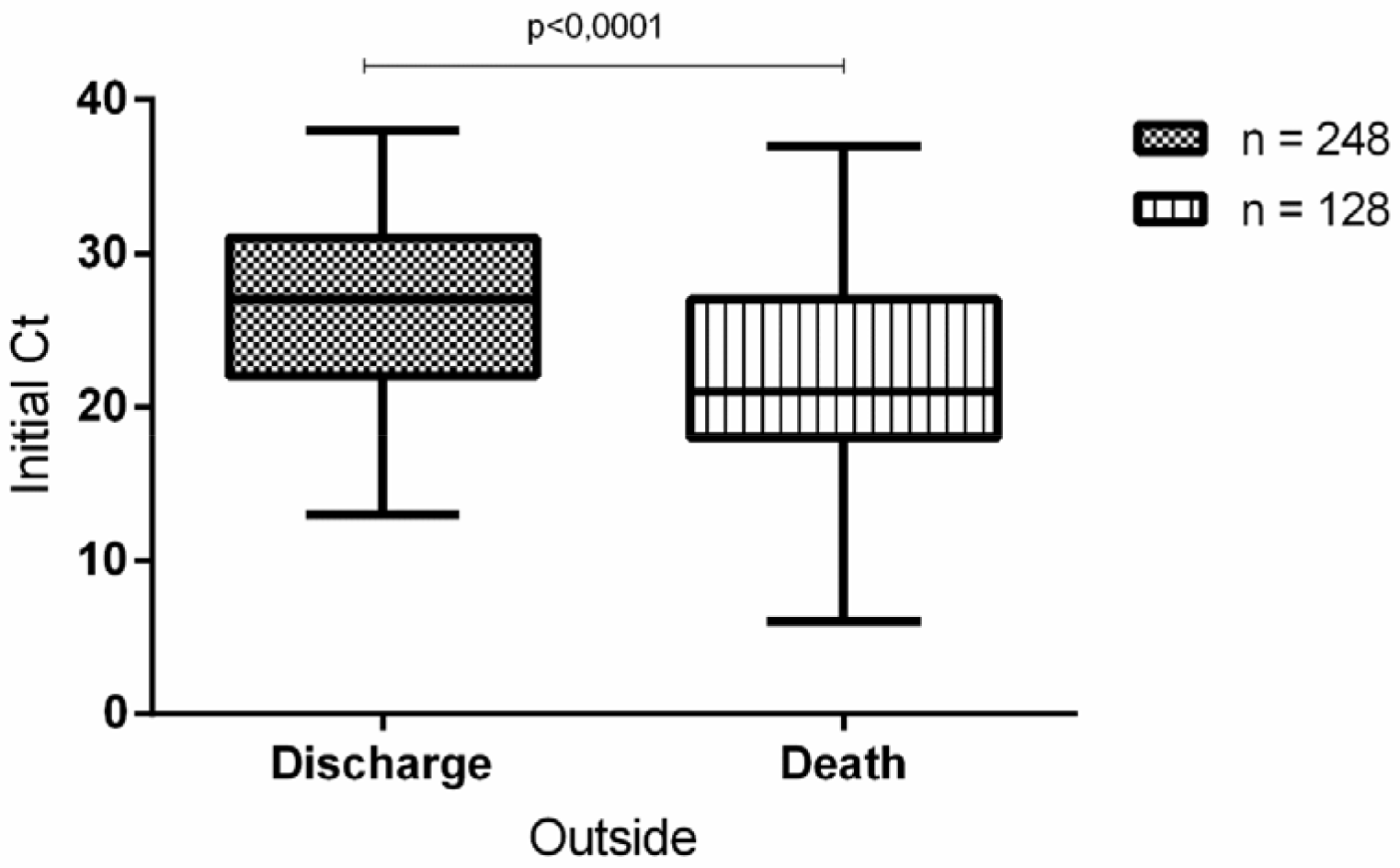
Ct values from the swab samples according discharge or death

## DISCUSSION

The analysis of viral load is applied in different endemic respiratory viruses’ diseases, although a clear association between the viral load and disease severity or outcome remains unelucidated. [11,12,13,14]. Nonetheless, certain viruses, such as H5N1, are considerably associated with severe disease, such as COVID-19. Pawestri et al. confirmed the association of the viral load with poor disease outcome.

Our study included a large number of admission SARS-CoV-2 samples with a stratified clinical expression in mild (non-hospitalized), moderate, and severe (hospitalized) than described by others who did not categorize patients.

Argyropoulos et al. [16] found that patients with mild Covid-19 infection presented high viral load, as was demonstrated in our study. However, their study demonstrated that hospitalized patients had a lower viral load than non-hospitalized patients, but they did not categorize the patients according to disease severity. Their small sample size probably accounts for the different results we observed in this study. Viral load distribution according to the Ct values from initial samples of our study patients followed a V curve, with mild and critically ill patients having higher viral load than hospitalized patients with a better outcome.

Ct values of <25 were consistent with high viral load in patients who were more likely to die during hospitalization. Magleby et al. [17] evaluating the correlation between Ct values and clinical outcome demonstrated a significant association between higher viral load and death or intubation.

The results of this study must be interpreted considering the methodological limitations. We focused on virologic aspects and did not include the effects of comorbidities, clinical symptoms, date of admission, date of sample collection, use of antivirals and antibiotics because of the heterogenous nature of the hospitalized patients and requirement of a large cohort for subgroup analysis. The duration of symptoms before testing may be an important variable, therefore, date of symptom onset is not consistently reported by medical assistants collecting samples at health services but other studies demonstrated that patients with other acute viral infections tend to present to hospital after expected peak viral load [18].

In conclusion, we found that admission SARS-CoV-2 viral load, as was determined by the Ct value, is an important surrogate epidemiological marker of infectivity that was independently associated with mortality among the patients hospitalized with COVID-19. These findings suggest that Ct values can be used to assist clinicians to identify patients at a high risk for severe outcome.

## Data Availability

none

